# Somatic Ras/Raf/MAPK Variants Enriched in the Hippocampus in Drug-Resistant Mesial Temporal Lobe Epilepsy

**DOI:** 10.1101/2022.12.23.22283854

**Authors:** Sattar Khoshkhoo, Yilan Wang, Yasmine Chahine, E. Zeynep Erson-Omay, Stephanie Robert, Emre Kiziltug, Eyiyemisi C. Damisah, Carol Nelson-Williams, Guangya Zhu, Wenna Kong, August Yue Huang, Edward Stronge, H. Westley Phillips, Brian H. Chhouk, Sara Bizzotto, Ming Hui Chen, Thiuni N. Adikari, Zimeng Ye, Tom Witkowski, Dulcie Lai, Nadine Lee, Julie Lokan, Ingrid E. Scheffer, Samuel F. Berkovic, Shozeb Haider, Michael S. Hildebrand, Edward Yang, Murat Gunel, Richard P. Lifton, R Mark Richardson, Ingmar Blümcke, Sanda Alexandrescu, Anita Huttner, Erin L. Heinzen, Jidong Zhu, Annapurna Poduri, Nihal DeLanerolle, Dennis D. Spencer, Eunjung Alice Lee, Christopher A. Walsh, Kristopher T. Kahle

**Affiliations:** Department of Neurology, Brigham and Women’s Hospital, Harvard Medical School, Boston, MA, USA; Division of Genetics and Genomics, Manton Center for Orphan Disease Research, Boston Children’s Hospital, Boston, MA, USA; Broad Institute of MIT and Harvard, Cambridge, MA, USA; Program in Biological and Biomedical Sciences, Harvard Medical School, Boston, MA, USA; Department of Neurosurgery, Yale University School of Medicine, New Haven, CT, USA; Department of Genetics, Yale University School of Medicine, New Haven, CT, USA; Interdisciplinary Research Center on Biology and Chemistry, Shanghai Institute of Organic Chemistry, Chinese Academy of Sciences, Shanghai 201203, China; Department of Pediatrics, Harvard Medical School, Boston, MA, USA; Program in Neuroscience, Harvard/MIT MD-PHD Program, Harvard Medical School, Boston, MA, USA; Department of Neurosurgery, David Geffen School of Medicine at University of California, Los Angeles, Los Angeles, CA, USA; Sorbonne University, Paris Brain Institute (ICM), National Institute of Health and Medical Research (INSERM), National Center for Scientific Research (CNRS), Paris, France; Department of Medicine (Austin Health), University of Melbourne, Heidelberg, Australia; Division of Pharmacotherapy and Experimental Therapeutics, Eshelman School of Pharmacy, University of North Carolina, Chapel Hill, North Carolina, USA; Department of Anatomical Pathology, Austin Health, Heidelberg, Australia; Murdoch Children’s Research Institute, Parkville, Australia; Florey Institute of Neuroscience and Mental Health, Heidelberg, Australia; Department of Pediatrics, University of Melbourne, Royal Children’s Hospital, Parkville, Australia; Bladin-Berkovic Comprehensive Epilepsy Program, Department of Neurology, Austin Health, Heidelberg, Australia; Department of Pharmaceutical and Biological Chemistry, University College London School of Pharmacy, London, UK; Department of Radiology, Boston Children’s Hospital, Harvard Medical School, Boston, MA, USA; Laboratory of Human Genetics and Genomics, The Rockefeller University, New York, NY, USA; Department of Neurosurgery, Massachusetts General Hospital, Boston, MA, USA; Department of Neuropathology, University Hospitals Erlangen, Erlangen, Germany; Epilepsy Center, Cleveland Clinic, Cleveland, OH, USA; Department of Pathology, Boston Children’s Hospital, Harvard Medical School, Boston, MA, USA; Department of Pathology, Yale University School of Medicine, New Haven, CT, USA; Department of Genetics, School of Medicine, University of North Carolina, Chapel Hill, NC, USA; Epilepsy Genetics Program, Division of Epilepsy and Neurophysiology, Department of Neurology, Boston Children’s Hospital, Harvard Medical School, Boston, MA, USA; Department of Neurology and Pediatrics, Harvard Medical School, Boston, MA, USA; Allen Discovery Center for Human Brain Evolution, Boston Children’s Hospital, Harvard Medical School, Boston, MA, USA; Howard Hughes Medical Institute, Boston, MA, USA; Department of Neurosurgery, Boston Children’s Hospital, Boston, MA, USA

## Abstract

**Importance:** Mesial temporal lobe epilepsy (MTLE) is the most common focal epilepsy subtype and is often refractory to anti-seizure medications. While most MTLE patients do not have pathogenic germline genetic variants, the contribution of post-zygotic (i.e., somatic) variants in the brain is unknown.

**Objective:** To test the association between pathogenic somatic variants in the hippocampus and MTLE.

**Design:** This case-control genetic association study analyzed the DNA derived from hippocampal tissue of neurosurgically-treated patients with MTLE and age- and sex-matched neurotypical controls. Participants were enrolled from 1988 through 2019 and clinical data was collected retrospectively. Whole-exome and gene-panel sequencing (depth>500X) were used to identify candidate pathogenic somatic variants. A subset of novel variants were functionally evaluated using cellular and molecular assays.

**Setting:** Level 4 epilepsy centers, multi-center study.

**Participants:** Non-lesional and lesional (mesial temporal sclerosis, focal cortical dysplasia, and low-grade epilepsy-associated tumors) drug-resistant MTLE patients who underwent anterior medial temporal lobectomy. All patients with available frozen tissue and appropriate consents were included. Control brain tissue was obtained from neurotypical donors at brain banks.

**Exposures:** Drug-resistant MTLE.

**Main Outcomes and Measures:** Presence and abundance of pathogenic somatic variants in the hippocampus versus the unaffected temporal neocortex.

**Results:** Samples were obtained from 105 MTLE patients (52 male, 53 female; age: MED [IQR], 32 [26-44]) and 30 neurotypical controls (19 male, 11 female; age: MED [IQR], 37 [18-53]). Eleven pathogenic somatic variants, enriched in the hippocampus relative to the unaffected temporal neocortex (MED [IQR], 1.92 [1.5-2.7] vs 0.3 [0-0.9], p<0.05), were detected in MTLE patients but not in the controls. Ten of these variants were in *PTPN11, SOS1, KRAS, BRAF*, and *NF1*, all predicted to constitutively activate Ras/Raf/MAPK signaling. Immunohistochemical studies of variant-positive hippocampal tissue demonstrated increased Erk1/2 phosphorylation, indicative of Ras/Raf/MAPK activation, predominantly in glial cells. Molecular assays showed abnormal liquid-liquid phase separation for the *PTPN11* variants as a possible dominant gain-of-function mechanism.

**Conclusions and Relevance:** Hippocampal somatic variants, particularly those activating Ras/Raf/MAPK signaling, may contribute to the pathogenesis of sporadic, drug-resistant MTLE. These findings may provide a novel genetic mechanism and highlight new therapeutic targets for this common indication for epilepsy surgery.

## Introduction

Epilepsy is a debilitating chronic neurologic condition that affects 1 in 26 people (3-4% lifetime risk)^1^. The most common focal epilepsy subtype, mesial temporal lobe epilepsy (MTLE), is often resistant to anti-seizure medications and requires neurosurgical intervention (e.g., anterior temporal lobectomy) in roughly one-third of the patients, with attendant morbidity^2^. While MTLE has been associated with initial precipitating insults such as prolonged febrile seizures^3^ and trauma^4^, its etiology is debated and its pathophysiology is poorly understood. With a few exceptions^5–8^, whole-exome sequencing (WES) and gene-panel sequencing (GPS) studies of blood- and buccal-derived DNA have had minimal success in identifying genetic determinants of focal epilepsies such as MTLE^9–14^, which typically occur in individuals without a family history of the disease.

Recently, post-zygotic (i.e., somatic) variants have emerged as a major cause of focal epilepsies associated with focal cortical dysplasia (FCD)^15^. Pathogenic somatic variants in FCD are present in only a fraction of cells (1-10% typically), creating a mosaic with admixed mutant (variant-positive) and non-mutant cells. The level of mosaicism, as defined by variant allele frequency (VAF), in focal epilepsy usually correlates with the size and brain regional distribution of the lesion^15^. Most somatic variants identified in FCD are seen in cases with type 2B histopathological classification and result in activation of PI3K/Akt/mTOR pathway genes that typically produce a lesion visible on MRI^15^. However, somatic variants in non-PI3K/Akt/mTOR genes can also cause focal epilepsies, without always producing a radiographically evident dysplasia, even at high VAFs^16^.

In this study, we tested the hypothesis that somatic variants enriched in the hippocampus are an important pathogenic mechanism underlying drug-resistant MTLE. We utilized deep WES and GPS to identify somatic variants in the hippocampal tissue from neurosurgically-treated MTLE patients and neurotypical controls. Here, we provide the first strong evidence of pathogenic somatic variants in MTLE and show that these variants activate the Ras/Raf/MAPK pathway—a clear distinction from the PI3K/Akt/mTOR pathway variants involved in extratemporal focal epilepsy. Additionally, we identified significant enrichment of these pathogenic variants in the affected hippocampus compared to the adjoining temporal neocortex, suggesting that the frequent success of mesial temporal resection or ablation in MTLE reflects the removal of tissue that is enriched for mutant cells. Our results define a new genetic pathway underlying focal epilepsy and highlight the importance of regional differences in cortical structure and development for models of epilepsy pathogenesis.

## Methods

### Study Participants

This case-control study was designed to investigate the association between somatic genetic variants and MTLE. Patients with non-lesional and lesional (mesial temporal sclerosis [MTS], focal cortical dysplasia, and low-grade epilepsy-associated tumors [LEATs: ganglioglioma, dysembryoplastic neuroepithelial tumor, and multinodular and vacuolating neuronal tumor]) drug-resistant MTLE who underwent anterior mesial temporal lobe resection were included. All non-LEAT primary brain tumors were excluded. Neurotypical control brain tissue was obtained from the NIH NeuroBioBank at the University of Maryland, Baltimore, and the University of Miami, and the European Epilepsy Brain Bank.

### Oversight and Study Procedures

The main study group comprised MTLE patients at Yale-New Haven Hospital, Boston Children’s Hospital, Austin Hospital, and the Royal Melbourne Hospital. Written informed consent was obtained from all patients or their guardians with local Institutional Review Board or Ethics Committee approval. All research performed with samples obtained from patients was approved by the Institutional Review Board at Boston Children’s Hospital. Participants IDs are assigned by the research team are not identifiable personal or clinical information.

### Target Capture, Sequencing, and Somatic Variant Analysis

WES and GPS (mean depth>500X) was performed on DNA extracted from fresh-frozen brain tissue or non-brain tissue to detect somatic mutations, which were independently validated with amplicon sequencing or droplet digital Polymerase Chain Reaction (ddPCR).

### Cellular and Molecular Studies

Cell lines were transfected with wild-type and mutant human gene constructs and underwent immunoblotting and liquid-liquid phase separation assays. Additional histopathology experiments were performed on archival paraffin-embedded tissue as needed.

### Statistical Analysis

Pairwise comparisons were made with Fisher’s exact test, Student’s t-test, Wilcoxon rank-sum and signed-rank tests, and the binomial test; p-values less than 0.05 were considered statistically significant. Pathway enrichment analysis^17^ and gene set over-representation analysis were evaluated using the hypergeometric test; False Discovery Rate (FDR)-adjusted p-values less than 0.05 were considered statistically significant.

Additional details are provided in Supplementary Methods.

## Results

### Patients

Surgical samples were obtained from 105 patients who underwent anterior medial temporal lobectomy for medication-refractory MTLE from 1988 through 2019. Clinical histopathologic findings included MTS-only (n=91), MTS+ (n=6; including LEATs [n=4] or FCD [n=2] in addition to MTS), non-MTS (n=8). Detailed clinical and demographic information is provided in Supplementary Table 1. All patients had hippocampal tissue available. A subset also had paired temporal neocortical tissue (n=89) and blood (n=27) collected. Only fresh-frozen specimens were employed for WES or GPS and validation.

### Pathogenic Somatic Variant Discovery and Brain Regional Enrichment Analysis

Hippocampus-derived DNA from patients underwent >500X WES or GPS (Supplementary Figure 1), followed by somatic variant calling. After careful bioinformatic measures to improve the accuracy of the somatic variant call set, all the variants previously reported as pathogenic or likely pathogenic in ClinVar^18^ were experimentally tested with amplicon sequencing and/or ddPCR. Of 21 candidate variants tested, 11 were validated. All validated pathogenic or likely pathogenic somatic variants detected in MTLE samples, and their corresponding VAFs, are shown in Table 1. Among the tissue samples with pathogenic somatic variants, 5 cases had MTS+ pathology with 2.5-23.6% mean VAFs. The remaining 6 cases with MTS-only pathology had significantly lower mean VAFs, ranging from 0.8 to 3.3% (p<0.01). We did not identify pathogenic somatic variants in the small non-MTS group.

**Table 1.**
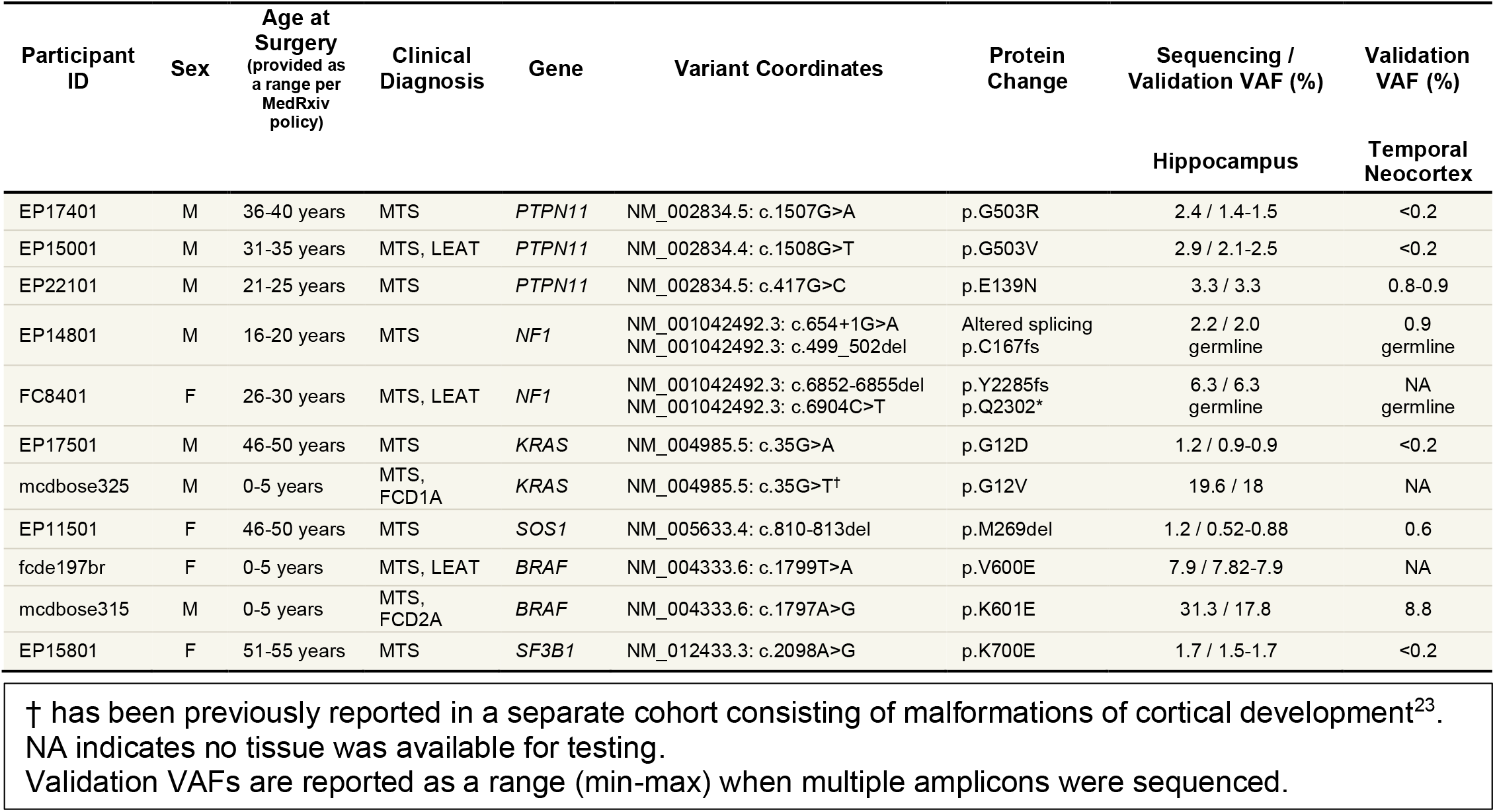
Pathogenic Somatic Variants in MTLE.

Since low VAF somatic variants may have been acquired at later developmental stages and be restricted to a small brain region, we also investigated the presence of pathogenic somatic variants in the unaffected temporal neocortex, when paired tissue was available. For all the variants that could be tested in both brain regions (n=8), the pathogenic variants were undetectable (<0.2% VAF, 4 patients) or less common (4 patients) in the temporal neocortex (Table 1), suggesting that variants were selectively enriched in affected hippocampal tissue (MED [IQR], 1.92 [1.5-2.7] vs 0.3 [0-0.9], p<0.05).

Notably, the *PTPN11* (c.1507G>A and c.1508G>T), *KRAS (*c.35G>A and c.35G>T) and *BRAF* (c.1797A>G and c.1799T>A) variants are all located in mutational hotspots for cancer and neurodevelopmental disorders^19–21^. However, except for *KRAS* c.35G>T and *BRAF* c.1799T>A which have been reported in FCDs^22,23^ and LEATs^24^, none of the other somatic variants have been previously described in focal epilepsies. Both patients with *NF1* somatic variants carried a diagnosis of neurofibromatosis type 1 and had known germline variants in the *NF1* gene (c.499_502del and c.6904C>T) based on clinical testing. Our findings of co-existent germline and somatic variants in MTLE patients are consistent with the established double-hit mechanism in *NF1*-associated pathology^25^.

To test whether pathogenic somatic variants exist in the hippocampus of neurotypical individuals, we performed WES on a cohort of 30 control hippocampal samples (Supplementary Figure 1). Since normal hippocampal tissue is rarely surgically resected, we used postmortem hippocampal dissections from individuals with no reported neurologic disease. In contrast to the MTLE cohort, we did not identify any pathogenic or likely pathogenic variants in the control samples.

### Clinical Findings in Patients with Activating Ras/Raf/MAPK Somatic Variants

All but one variant (SF3B1 p.K700E) in Table 1 are present in Ras/Raf/MAPK pathway genes (Figure 1A). Notably, all the variants are predicted to increase Ras/Raf/MAPK signaling via gain-of-function of a pathway activator (*PTPN11, KRAS, SOS1, BRAF*) or loss-of-function of a pathway repressor (*NF1*). To determine whether patients with activating Ras/Raf/MAPK variants have unique clinical characteristics, we independently reviewed all available clinical data, MRI images, and histopathology slides for this subgroup. All variant-positive patients were seizure-free >2 years after surgery (Engel class IA / ILAE class 1), with significantly increased likelihood of Engel class IA outcome (p<0.05) compared to the rest of the cohort (Supplementary Figure 2). All the patients with pathogenic Ras/Raf/MAPK variants had evidence of MTS; in 5 patients that was the only major imaging and histopathologic finding (Supplementary Figure 3A-P) while the remaining half exhibited additional findings consistent with FCDs or LEATs (Supplementary Figure 4). To further rule out the possibility of radiographically and histopathologically undetected LEATs, we performed additional CD34 staining—a diagnostic marker for LEATs^24^—in 4 samples with MTS-only pathology for which tissue was available. We did not observe higher than background-level CD34 staining (Supplementary Figure 3M-P), further confirming that Ras/Raf/MAPK variants can be present at low VAFs in the affected temporal lobe in the absence of a glioneuronal tumor detected radiographically or histopathologically.

**Figure 1.**
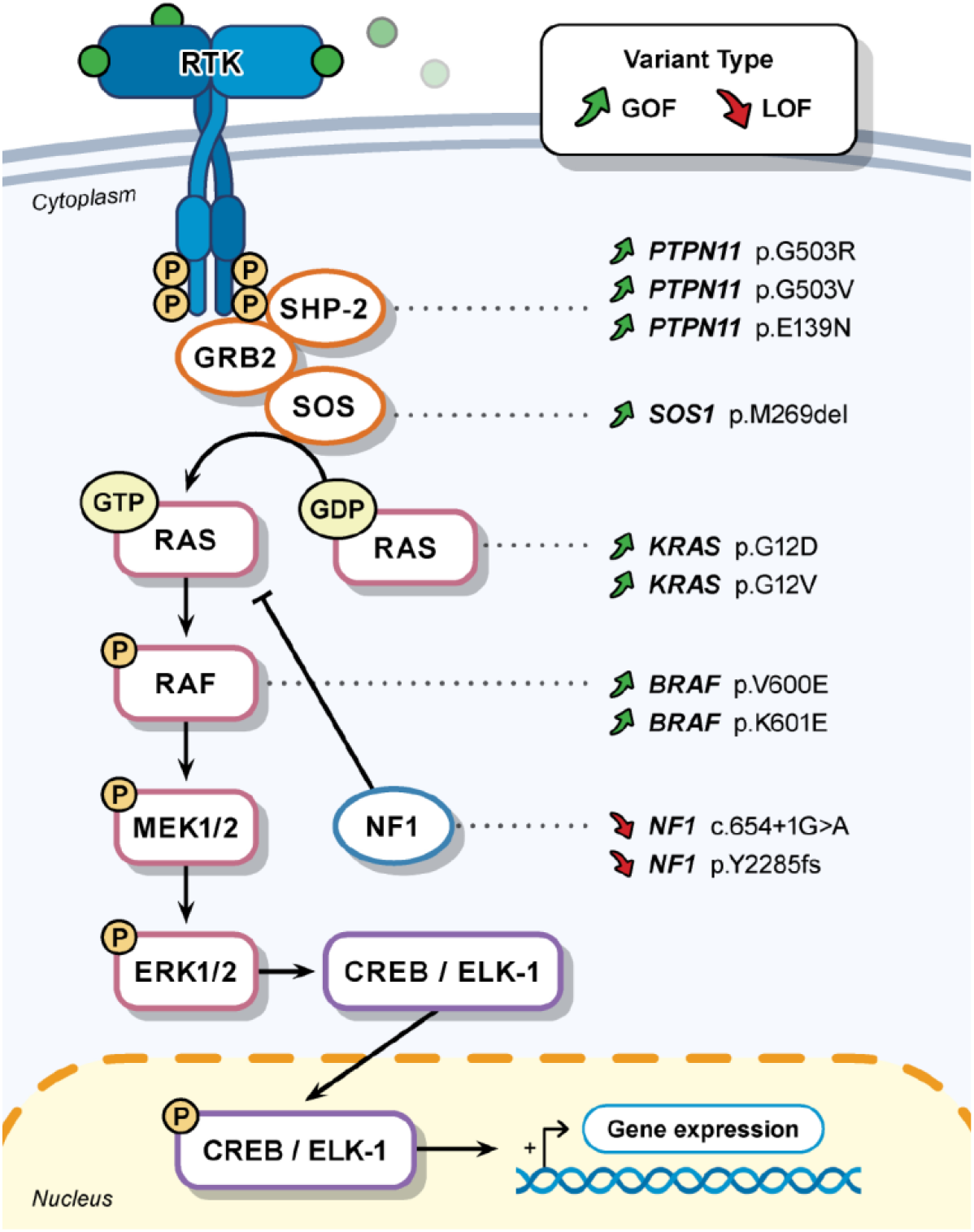
Somatic Variants Activating Ras/Raf/MAPK Pathway Genes in MTLE. Simplified diagram of the Ras/Raf/MAPK signaling pathway, the pathogenic variants discovered in the MTLE cohort, and their corresponding proteins. RTK: Receptor Tyrosine Kinase.

### Enrichment of Activating Ras/Raf/MAPK Pathway Variants in MTLE

While the contribution of somatic variants to MTLE has not been previously described, most FCD-associated somatic variants reported to date are in PI3K/Akt/mTOR pathway genes^15^ whereas LEAT-associated variants often involve the Ras/Raf/MAPK pathway^24^. To further investigate the relationship between somatic genotype and brain regional specificity, we performed a focused retrospective review of the FCD and LEAT literature. Our analysis demonstrated a significant predilection of somatic Ras/Raf/MAPK variants for the temporal lobe, while PI3K/Akt/mTOR variants showed an extra-temporal predominance (Figure 2A, p<0.001). Given that the hippocampus is the primary affected region in MTLE, our findings lend further support to the hypothesis that somatic variants in the Ras/Raf/MAPK pathway arising specifically in the temporal lobe confer risk for focal epilepsy.

**Figure 2.**
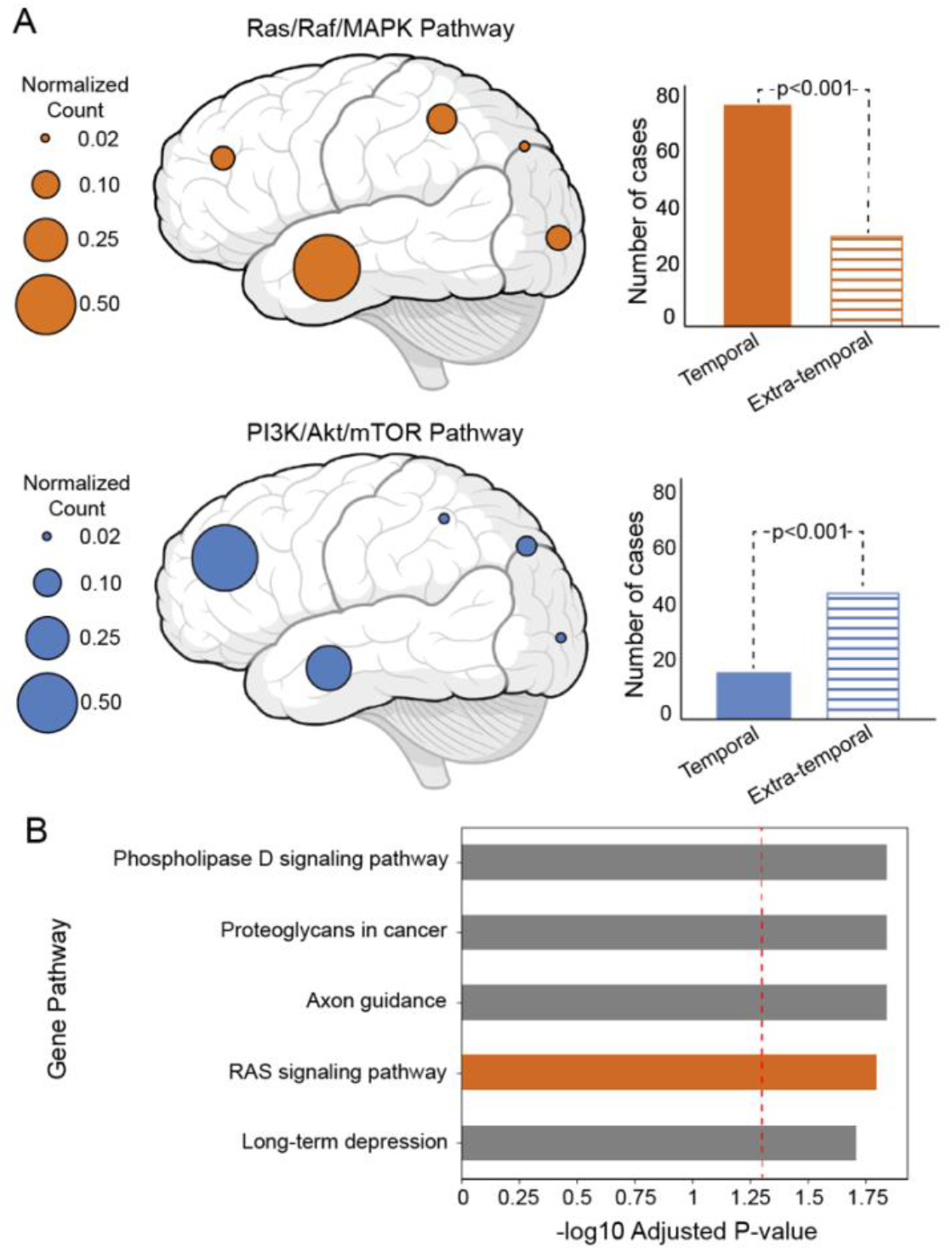
Enrichment of Ras/Raf/MAPK Pathway Variants in the Temporal Lobe and in MTLE. A) Retrospective review of Ras/Raf/MAPK pathway variants and PI3K/Akt/mTOR pathway variants in the lesional focal epilepsy literature. Circle diameters represent normalized case counts in each brain region and the associated bar plots on the right depict the absolute number of cases. B) KEGG (Kyoto Encyclopedia of Genes and Genomes) pathway enrichment analysis on the pathogenicity enriched variants in the MTLE cohort (p-value<0.001 and adjusted p-value<0.05 after correction for multiple hypotheses testing). The dashed red line represents the adjusted p-value of 0.05.

Since our initial detection of pathogenic somatic variants relied on prior reporting of a specific variant in ClinVar, we next performed pathway enrichment analysis on all the filtered variants from our call set that were predicted to be pathogenic (Supplementary Table 2). Consistent with our initial observation, for somatic variants in the MTLE cohort the Ras/Raf/MAPK signaling pathway was amongst the most highly enriched pathways (Figure 2B, adjusted p<0.05). Enrichment in this pathway was not seen in the control samples. Furthermore, we showed that pathogenicity-enriched variants in another curated Ras/Raf/MAPK gene set (Supplementary Table 3) are over-represented in the MTLE cohort (p<0.001) and not the control samples (p=1). Notably, we did not see an over-representation of PI3K/Akt/mTOR genes (Supplementary Table 3) in the MTLE cohort (p=1), further supporting the notion that focal epilepsies in the temporal lobe share a genetic etiology distinct from focal extra-temporal epilepsies.

### Mechanism of Ras/Raf/MAPK Overactivation in MTLE-associated *PTPN11* Somatic Variants

Germline *PTPN11* variants which are known causes of Noonan syndrome and related disorders appear to enhance protein tyrosine phosphatase enzymatic activity through an acquired capability of liquid-liquid phase separation (LLPS) of the mutant Shp2 protein encoded by the *PTPN11* variants^26^. Since this gene has not been previously associated with focal epilepsy, we evaluated the functional impact of MTLE-associated *PTPN11* variants by transiently expressing the human wild type (Shp2^wt^) and mutant (Shp2^mut^) constructs in HEK293T cells and then performing immunoblotting to assess the degree of Erk1/2 (a downstream effector of the Ras/Raf/MAPK pathway) phosphorylation. Compared to cells expressing the Shp2^wt^ protein, we detected increased phosphorylated Erk1/2 (pErk1/2) in cells expressing Shp2^mut^ proteins (Figure 3A), indicating increased phosphatase activity of all the Shp2 variants consistent with constitutive activation.

**Figure 3.**
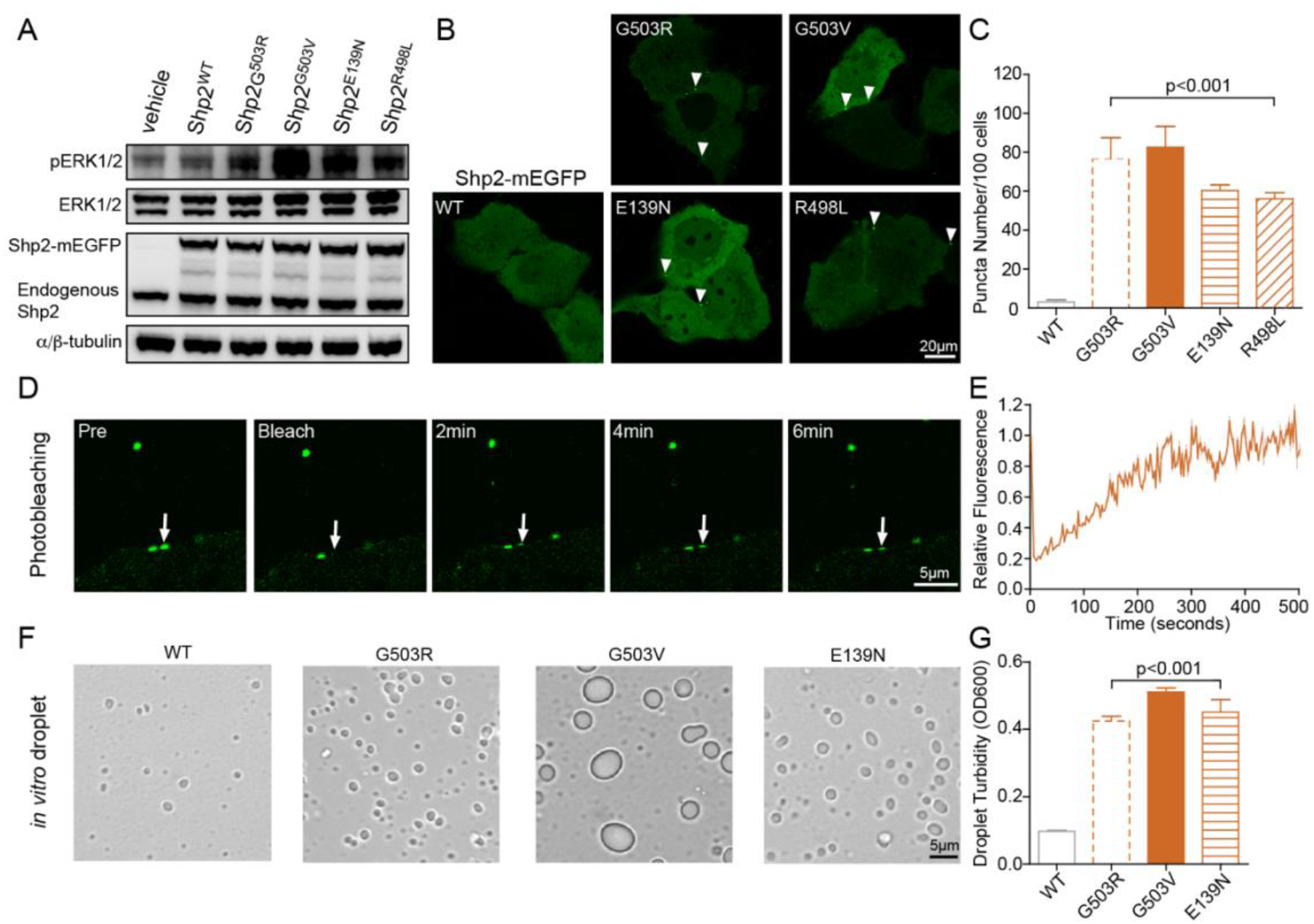
Mechanisms of Ras/Raf/MAPK Overactivity in MTLE-Associated Mutant Shp2 Proteins. A) Immunoblot of the indicated proteins in HEK293T cells transiently expressing Shp2^wt^ and Shp2^mut^. B) Representative pictures from live imaging microscopy of Shp2^wt^-mEGFP and Shp2^mut^-mEGFP proteins, transiently expressed in KYSE520 cells. The fluorescent puncta (marked by white arrowheads) are quantified in panel C (mean±SEM, p<0.001 mut vs wt). D) Representative images of FRAP using the transiently expressed Shp2^G503R^-mEGFP protein in KYSE520 cells. The rate of fluorescence recovery is quantified in panel E (mean±SEM, n=3 experiments). F) Representative images of 8µM Shp2^wt^ and Shp2^mut^ protein droplet formation in the presence of 10% (w/v) PEG3350 *in vitro*. Droplet formation is quantified by solution turbidity of OD600 in panel G (mean±SEM, p<0.001 mut vs wt).

To determine the mechanism through which MTLE-associated *PTPN11* variants enhance phosphatase activity, we tested whether these variants also undergo LLPS. We transiently expressed mEGFP-labeled Shp2 proteins (WT, G503R, G503V, E139N, R498L) in KYSE520 cells. Remarkably, the mutant Shp2 variants formed discrete puncta in cells, whereas the wild-type protein was dispersed throughout the cell (Figure 3B-C). Fluorescence recovery after photobleaching (FRAP) experiments showed that Shp2^G503R^-mEGFP puncta recovered within minutes upon photobleaching (Figure 3D-E), indicating that the mutant Shp2 proteins formed condensates that exhibited dynamic liquid-like behavior. To explore whether mutant Shp2 proteins could also undergo LLPS *in vitro*, we expressed and purified recombinant Shp2 mutant and wild-type proteins. The *in vitro* droplet formation assay^26^ showed that the mutant Shp2 proteins formed more and larger droplets compared to the wild-type protein in the presence of 10% (w/v) PEG3350, which was quantified using solution turbidity of OD600 (Figure 3F-G). These findings indicate that the MTLE-associated mutant Shp2 proteins undergo LLPS in cells and *in vitro* through a dominant gain-of-function mechanism that activates downstream Ras/Raf/MAPK signaling.

### Ras/Raf/MAPK Overactivation in Affected Human MTLE Surgical Tissue

Given the enrichment of activating Ras/Raf/MAPK somatic variants in MTLE, the variant-positive cells could play a key and potentially causal role in hippocampal epileptogenesis. In the absence of allele-specific antibodies for most of the variants found in our cohort, we used immunohistochemical staining for pErk1/2 (a proxy for Ras/Raf/MAPK pathway overactivation^27^) to determine the identity and localization of variant-positive cells. In brain tissue from the LEAT case associated with *PTPN11* p.G503V, we observed substantial correlation between tumor density (Figure 4A) and the degree of pErk1/2 staining (Figure 4F), supporting the validity of this strategy in localizing variant-positive cells. Using this approach in two cases with NF1 c.654+1G>A and KRAS p.G12D variants and MTS-only pathology, we observed that hippocampal subregions with the greatest neuronal loss showed the highest density of pErk1/2 staining (Figure 4B-E, H-I). Most cells with intense pErk1/2 staining in areas of neuronal loss showed glial morphology (Figure 4G, H1, I), which could not be explained by the differences in cell type-specific gene expression in the adult human hippocampus (Supplementary Figure 5).

**Figure 4.**
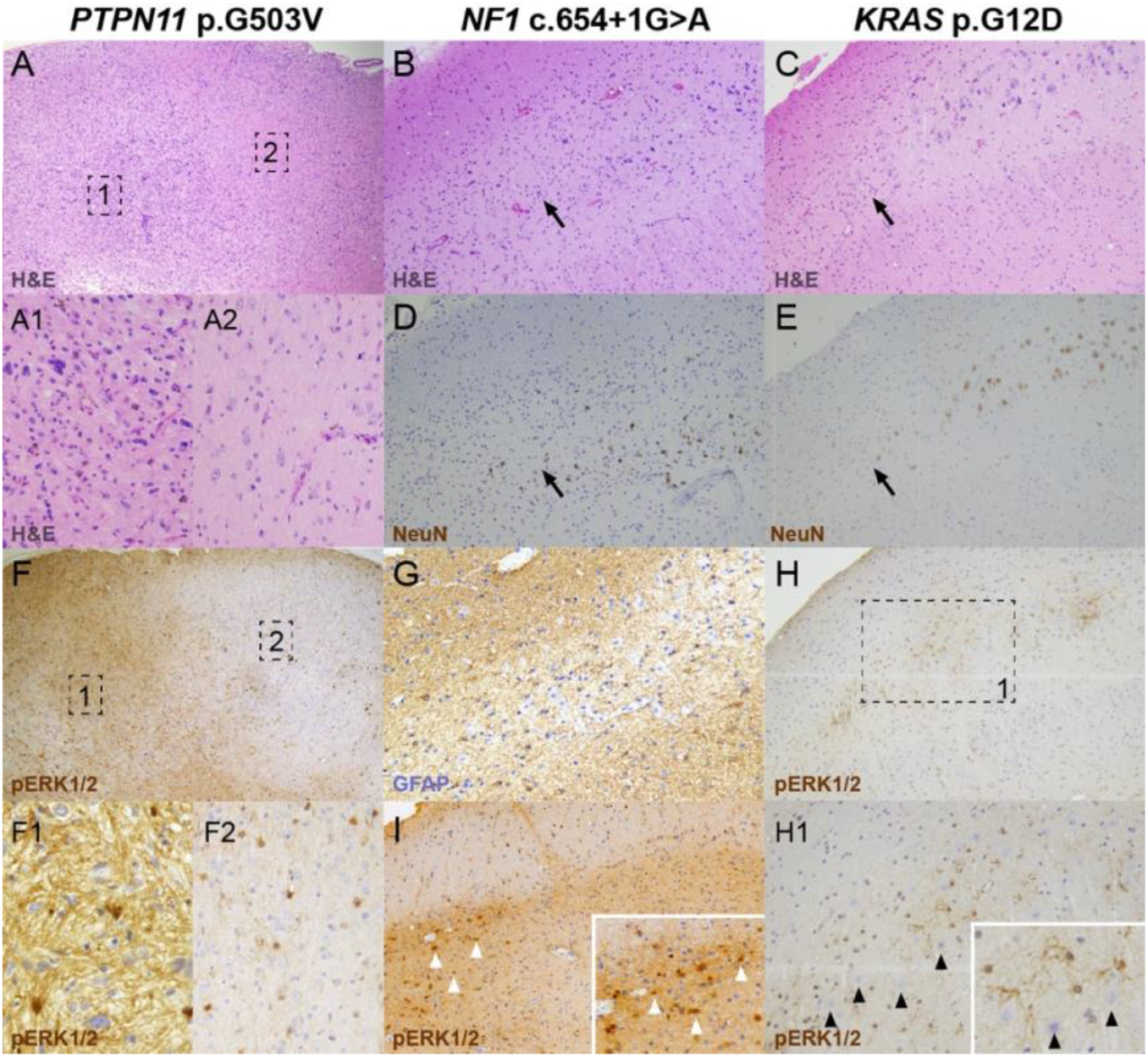
Evidence of Erk1/2 Overactivation in MTLE Tissue Harboring Ras/Raf/MAPK Variants. Each column shows histopathologic images from one sample. A, F) A LEAT with areas of hypercellularity corresponding to the tumor (A1) intermixed with normal brain tissue (A2). pErk1/2 staining of the corresponding regions is seen in F, F1-2. B-E, G-H) Representative MTS-only samples with neuronal dropout in the Cornu Ammonis (CA) region of the hippocampus (B-E) but uniform distribution of glial cells across the same region (G). The black arrows demarcate CA neuronal loss. Erk1/2 phosphorylation colocalizes to regions with highest degree of neuronal loss and sclerosis (H, I). The white arrowheads point to cells with increased pErk1/2 staining, mostly represented by apparent glia as highlighted in the panel I inset. The black arrowheads point to neurons which have relatively low pErk1/2 staining as highlighted in the panel H1 inset. The higher resolution images were obtained at 200-400X and the lower resolution images were obtained at 40X.

## Discussion

We identified pathogenic somatic variants enriched in the hippocampus, where seizures in MTLE typically originate, in 11 patients with drug-resistant MTLE, with none found in the neurotypical controls. Strikingly, all but one of these variants are predicted to activate Ras/Raf/MAPK signaling, providing strong evidence that they may contribute to MTLE risk, analogous to the role of somatic PI3K/Akt/mTOR variants in FCD. The finding that even MTLE cases without evidence of dysplasia or neoplasia carry pathogenic somatic variants in the Ras/Raf/MAPK pathway could significantly alter our understanding of, and the therapeutic options for, this most common indication for epilepsy surgery.

The regional specificity of epilepsy-associated Ras/Raf/MAPK pathway variants in the temporal lobe, versus PI3K/Akt/mTOR pathway variants in the extra-temporal cortex, recalls cancers where common driver mutations are characteristic to particular cell and tissue types^19^. Hence the regional genetic specificity of focal epilepsies may represent the major differences in cellular architecture and proliferative properties between different brain regions. The higher VAFs of Ras/Raf/MAPK variants in the hippocampus compared to the lateral temporal neocortex parallels the pattern of ongoing neurogliogenesis, which continues after birth only in the dentate gyrus^28^, and may contribute to this enrichment. Febrile seizures and head trauma, well-established risk factors for MTLE^3,4^, stimulate this dentate gyrus proliferation in animal models^29,30^. A similar process in humans could create a proliferative or survival advantage for cells harboring activating Ras/Raf/MAPK pathway variants, as has been described in cancer^31^, and could provide a potential mechanism for how these risk factors may contribute to MTLE risk.

We also observed a significant association between the lower VAF of pathogenic variants and the absence of dysplasia or neoplasia on imaging and histopathology. This finding implies that patients with MTS-only pathology may have acquired their variants at later developmental stages^15^, and therefore have fewer variant-positive cells. Patients with MTS+ pathology, in contrast, may have acquired their variants earlier resulting in higher fractions of variant-positive cells and diagnosis of drug-resistant epilepsy at a younger age, as illustrated by mcdbose325 and mcdbose315 who underwent epilepsy surgery as infants. Since most patients in our cohort are adults and due to limitations of retrospective access to clinical information, we were unable to test the association between the age of onset or epilepsy duration with VAF. In addition to VAF, other factors may contribute to the histopathology associated with Ras/Ras/MAPK variants. For example, EP17401 and EP15001 both have *PTPN11* variants with similar VAFs that change the same amino acid, but a greater degree of Ras-MAPK activation by *PTPN11* p.G503V may account for MTS+ pathology as opposed to MTS-only with p.G503R. The cellular lineage of variant-positive cells is likely an independent predictor of pathology as well^32^. Consequently, somatic Ras/Raf/MAPK pathway variants may give rise to a spectrum of temporal lobe lesions depending on the developmental timepoint at which they were acquired, their molecular mechanisms, and the cell types of variant-positive cells, all associated with drug-resistant MTLE.

The mechanism through which somatic Ras/Raf/MAPK variants contribute to epileptogenesis is yet to be determined, although our functional data support the role of Ras/Raf/MAPK overactivation in this process. While all variant-positive patients exhibited histopathologic evidence of MTS, we did not identify pathogenic somatic variants in most patients with MTS, and very few patients with non-MTS histopathology were enrolled in the study; therefore, the exact relationship between MTS and pathogenic somatic variants remains unclear based on our data. The precise cell type-specific effects of somatic Ras/Raf/MAPK pathway variants in MTLE are likely quite complicated and may evolve over time, given that these variants can cause different phenotypes in distinct cell types, contexts, and brain regions^33,34^. Our human genetic findings provide the impetus to explore these hypotheses in future work with animal models.

Our study highlights the importance of incorporating molecular testing into the MTLE diagnostic algorithm. Although it is not known yet whether MTLE-associated somatic variants can be detected less invasively, perhaps presurgical evaluation of cell-free DNA in CSF^35,36^ or genomic DNA in SEEG electrodes^37^, could guide further management steps such as surgical approaches versus genotype-driven therapeutics. For example, in patients with left MTLE who may be at risk of significant verbal memory or language decline with an extensive resection, or in patients who do not receive surgery due to patient/provider perception^38^ or limited resources^39^, a genotype-driven pharmacological approach may provide an additional treatment option. Since many targeted treatments for Ras/Raf/MAPK activation are already in various stages of clinical testing in cancers^40^, our findings offer the potential to leverage some of these agents to develop the first generation of targeted therapies for molecularly characterized MTLE.

### Limitations

Despite the high sequencing depth, somatic variant detection was mostly limited to VAFs>1% due to technical factors. Since cell loss is a prominent feature of MTS, pathogenic variants in the hippocampus may be present at VAFs<1%, suggesting that the burden of pathogenic somatic variants in MTLE could be much higher than what is reported here.

## Data Availability

All the whole-exome and gene-panel sequencing data generated for this study will be deposited in the database of Genotypes and Phenotypes (dbGaP).

## Conclusions and Relevance

Hippocampal somatic variants, particularly those activating Ras/Raf/MAPK signaling, may contribute to the pathogenesis of sporadic, drug-resistant MTLE. These findings may provide a novel genetic mechanism and highlight new therapeutic targets for this common indication for epilepsy surgery.

## Data Sharing

All the WES and GPS data generated for this study will be deposited in the database of Genotypes and Phenotypes (dbGaP) at the time of publication and may be accessed through the following accession numbers: phs000492.v4.p2 and phs002128.v1.

## Author Contributions

SK, IB, EK, IES, SFB, AP, SR, ED, ND, and DS enrolled the subjects and collected tissue for sequencing. SK, SR, IES, SFB, MHC, AP, and DS performed clinical phenotyping. JL, SA, and AH completed the initial histopathologic review/classification and performed additional staining of paraffin-embedded tissue slides as needed. IB performed an additional independent review of the histopathologic findings. EY performed an additional independent review of all the radiologic findings. SK, ES, HWP, SB, TA, ZY, TW, DL, MSH, ELH, KTK, and CAW performed sequencing experiments. SK, YW, AYH, EZE, DL, ELH, EAL, KTK, and CAW performed DNA sequencing analysis and variant discovery. SK, YC, BHC, TA, ZY, TW, DL, and MSH performed variant validation experiments. SK, SR, GZ, WK, JZ, AH, CNW, KTK performed the molecular phenotyping experiments and SH performed the biophysical modeling of mutations. SK and NL performed a retrospective review of the focal epilepsy literature. SK, YW, AYH, EAL performed the statistical analyses. SK, YW, EAL, MG, RPL, KTK, and CAW wrote the manuscript. SK and KTK had full access to all the data in the study and take responsibility for the integrity of the data and the accuracy of the data analysis.

### Other Contributors

We would like to thank the following individuals for their valuable contributions including assistance with obtaining brain tissue for research, discussion of experimental findings, and comments on the manuscript: Zinan Zhou, PhD (Boston Children’s Hospital), Michael B. Miller, MD, PhD (Brigham and Women’s Hospital), Javier Ganz, PhD (Boston Children’s Hospital), Shyam Akula, PhD (Harvard Medical School), Abbe Lai, MS, CGC (Boston Children’s Hospital), Jennifer E. Neil, MS, CGC (Boston Children’s Hospital).

