## Supplementary Information for "Somatic Ras/Raf/MAPK Variants Enriched in the Hippocampus in Drug-Resistant Mesial Temporal Lobe Epilepsy"

### Supplementary Methods

Confirmation of Clinical, Radiologic, and Histopathologic Findings
Clinical and surgical information was obtained from patients' charts by the authorized study team. All diagnoses were confirmed by the study pathologists, radiologists, and neurosurgeons. Radiology images and histopathology slides for patients with tissue harboring pathogenic somatic variants were additionally reviewed by an independent expert neuroradiologist (EY) and neuropathologist (IB), respectively.

Brain Tissue Procurement
Brain tissue was labeled as hippocampus or temporal neocortex in the OR and then evaluted and allocated for research by the clincal neuropathology team. While in all cases anterior medial temporal tissue and anterior temporal neocortical tissue were collected, there may be some variability in the exact location that was sampled for genoimc DNA extraction.

Genomic DNA Extraction
Genomic DNA from fresh-frozen surgical samples and whole-blood was extracted using Chemagic DNA Tissue 100 H24 kit (Perkin Elmer), DNeasy Blood & Tissue kits (Qiagen), All Prep DNA/RNA kit (Qiagen), QIAamp DNA Blood kits (Qiagen), or a standard phenol chloroform extraction[1]. Standard quality and purity assessments were conducted via determination of the 260/280nm ratios for values of 1.7-2.0. All the genomic DNA samples were quantified using Qubit or PicoGreen (ThermoFisher Scientific) for proper assessment of double stranded DNA concentration.

Whole-Exome Sequencing
Genomic DNA derived from brain and non-brain tissue (Supplementary Table 1) underwent whole-exome capture using IDT xGen Exome Research Panel v1+custom spike-in probes (IDTERPv1custom), IDT xGen Exome Research Panel v1+mitochondrial genome spike-in probes (IDTERPv1mtDNA), or Twist Human Comprehensive Exome v1 (TwistHCEv1). For the samples prepared using the Twist capture probes, IDT xGen UDI-UMI adapters were substituted in to improve variant calling accuracy. The prepared paired-end whole-exome libraries were sequenced on NovaSeq 6000 at Yale Center for Genome Analysis, Harvard Biopolymers Facility, or Columbia University Irving Medical Center.

Gene-Panel Sequencing
Genomic DNA derived from brain tissue (Supplementary Table 1) underwent gene-panel sequencing. Starting from recurrent cancer hotspot mutations in a commercially-available panel (Cancer Hotspot v2, Thermo Fisher) and cancer-linked mutations in MTOR-pathway genes (COSMIC, accessed Nov. 2018), we selected gain-of-function SNPs, prioritizing variants occurring multiple times in COSMIC datasets and submitting top variants for Ampliseq pool design with manufacturer-designed software (Ampliseq, Thermo Fisher). To generate Ampliseq libraries, 10 ng of genomic DNA was added to 1X final Phusion U Hot Start Master Mix (Thermo Fisher), 0.9X Ampliseq primer pool, and 80 ng ET SSB (NEB), undergoing 17 cycles of PCR before standard Illumina library preparation. The panel was sequenced on a Nextseq 500 high output flow cell at Harvard Biopolymers Facility.

Alignment and Preprocessing of Sequencing Data
*IDT Whole-Exome Sequencing*
All the whole-exome sequencing data from cases and controls were aligned to hg38 reference genome (GRCh38.p13) using Burrows-Wheeler Aligner (BWA, version 0.7.17)[2]. We used the GATK Best Practices Workflow[3] for the remainder of preprocessing steps. Briefly, duplicates were marked using Picard[4] (version 2.18.14) MarkDuplicates. We then sorted and fixed the bam tags and performed base quality recalibration using GATK[3] (version 4.1.9.0) SortSam, SetNmMdAndUqTags, BaseRecalibrator, and ApplyBQSR.

*Twist Whole-Exome Sequencing*
Following the best practices for data analysis when using UMI adapters to improve variant detection[5], we demultiplexed the resulting binary base call (bcl) file for each sample and converted it to an unaligned bam file with UMI information using Picard (version 2.18.14) ExtractIlluminaBarcodes and IlluminaBasecallsToSam

functions. We then aligned the reads from the unaligned bam file to the hg38 reference genome (GRCh38.p13) and included the UMI tags from unmapped reads using Picard SamToFastq and MergeBamAlignment, BWA (version 0.7.17), and Samtools[6,7] (version 1.19) merge functions. The reads were then grouped by UMIs with the fgbio[8] (version 1.4.0) GroupReadsByUmi function. We then generated the consensus reads (n=1) generated with fgbio[8] CallMolecularConsensusReads function, sorted them with Picard SortSam, realigned them using BWA, and included the UMI tags from unmapped reads using Picard MergeBamAlignment. Lastly, we filtered the resulting consensus bam files with fgbio[8] FilterConsensusReads (min-reads=1, min-base-quality=20, max-no-call-fraction=0.05, min-mean-base-quality=10). To produce high-quality variant calls, we next performed additional preprocessing steps on the filtered consensus bam files. First, we sorted and fixed the bam tags using GATK (version 4.1.9.0) SortSam and SetNmMdAndUqTags. Second, we performed local indel realignment with GATK (version 3.8.1.0) RealignerTargetCreator and IndelRealigner. Last, we performed base quality score recalibration with GATK (version 4.1.9.0) BaseRecalibrator and ApplyBQSR.

*Custom Gene-Panel Sequencing*
All the whole-exome sequencing data from cases and controls were aligned to hg38 reference genome (GRCh38.p13) using BWA (version 0.7.17)[2]. We then sorted and fixed the bam tags and performed base quality recalibration using GATK (version 4.1.9.0) SortSam, SetNmMdAndUqTags, BaseRecalibrator, and ApplyBQSR.

<u>Somatic Single Nucleotide Variant (SNV) and Small Insertion-Deletion Variant (INDEL) Calling</u>
*Unpaired Calling*
Unpaired somatic variant calling was performed using GATK (version 4.1.9.0) MuTect2[9] using the hippocampal preprocessed bam files as “tumor”. The raw MuTect2[9] vcf files were filtered using GATK (version 4.1.9.0) FilterMutectCalls.

*Paired Calling*
For all the hippocampal samples with paired non-brain or neocortical tissue, the preprocessed bam files for the paired tissue were downsampled to 50X using sambamba[10] (version 0.7.1), mosdepth[11] (version 0.3.2), and customized python scripts. The downsampling was performed in order to minimize the chance of filtering out a shared somatic variant present in both the hippocampus and a paired tissue. Then paired somatic variant calling was performed with GATK (version 4.1.9.0) MuTect2[9] and Strelka2[12] (version 2.9) using the hippocampal bam files as “tumor” and the downsampled paired tissue bam files as “normal”. The raw MuTect2[9] vcf files were filtered using GATK (version 4.1.9.0) FilterMutectCalls. Finally, the filtered MuTect2 and Strelka2 call sets were merged to create a union call set with high sensitivity for somatic variant discovery.

*Paired Calling with a Panel of Normals (PON)*
For pathway enrichment analysis where accuracy was prioritized over sensitivity, a PON approach was used for variant calling to eliminate the false positive somatic calls caused by technical factors and error-prone genomic regions. Following a previously published strategy[13], for each case we created a unique PON using all the somatic variant call sets in the same cohort excluding the case that was being evaluated. This was achieved using GATK (version 4.1.9.0) GenomicDBImport followed by CreateSomaticPanelofNormals. Then we performed paired somatic variant calling using GATK (version 4.1.9.0) MuTect2[9] as described above, this time including a PON as an additional input. The raw MuTect2[9] vcf file was filtered by GATK (version 4.1.9.0) FilterMutectCalls. Since somatic variant calling could be affected by sequencing depth, for Gene Set Overrepresentation Analysis (described below) where cases and controls were directly compared side by side, the preprocessed bam files for the MTLE hippocampal tissue were downsampled to 284X to achieve the same mean sequencing depth for MTLE and control bam files.

<u>Somatic Variant Annotation</u>
All the variants were annotated using snpEff[14] (version 5.1). The databases included were the latest versions of gnomAD[15], ExAC[16], ClinVar[17], Rare Exome Variant Ensemble Learner (REVEL)[18], SNVs in Evolutionary model of Variant Effect (EVE)[19], Catalogue Of Somatic Mutations In Cancer (COSMIC, v95)[20], and SpliceAI[21] (version 1.3.1).

Pathogenic Somatic Variant Discovery

Since establishing pathogenicity in somatic variants can be challenging and frequently requires extensive functional validation, we limited the initial pathogenic variant discovery to known variants that were previously reported in ClinVar as "pathogenic" or "likely pathogenic." Only variants in genes with expression in the hippocampus based on The Genotype-Tissue Expression (GTEx) Project[22] were included in further analysis. Given the enrichment of Ras/Raf/MAPK variants based on the initial analysis, the raw (unfiltered) MuTect2 call sets were also independently screened for the presence of pathogenic or likely pathogenic Ras/Raf/MAPK variants. To remove the likely false positives, candidate variants were visually inspected using Integrative Genomics Viewer (IGV)[23] and only variants that met the following criteria were submitted for experimental confirmation:

1) Variant Allele Frequency (VAF) <0.3
2) Presence on at least 2 alignments with different start positions
3) Minimum 3 alternative (ALT) reads on forward and reverse orientations
4) Absence from error-prone genomic regions with multiple other somatic variants within a 50bp window.

Experimental confirmation was performed using amplicon sequencing or ddPCR as discussed separately.

Pairwise Statistical Tests of VAFs for Pathogenic Ras/Raf/MAPK Variants

We conducted pairwise comparisons of VAFs for pathogenic Ras/Raf/MAPK in:

1) Cases with MTS versus cases with MTS+ pathology (Two-sided Wilcoxon rank-sum test)
2) Cases with both hippocampal and temporal neocortical tissue (Two-sided Wilcoxon signed-rank test)

To determine the appropriate statistical test, for each pair of VAFs, we first tested whether they were normally distributed using the Shapiro test. We next tested the equality of variances of the pair (homoscedasticity). If both were normally distributed (Shapiro test p-value >= 0.05), we used the Bartlett test; if at least one was not normally distributed (Shapiro test p-value < 0.05), we used the Levene test. Last, we tested the equality of mean based on the previous tests of normality and homoscedasticity. If the pair passed both tests (Bartlett test p-value >= 0.05), we used Student's t-test; if the pair failed either test (Shapiro test p-value < 0.05 or Bartlett test p-value < 0.05 or Levene test p-value < 0.05), we used the Wilcoxon rank-sum test. In both scenarios, the pair was determined to be significantly different if p-value < 0.05. All statistical tests were implemented using the Python SciPy library[24].

Pairwise Statistical Test for Surgical Outcome

To determine whether the 2-year surgical outcome is different for the Ras/Ras/MAPK variant-positive patients compared to the rest of the cohort, we only included cases with sufficient clinical information at the 2-year postsurgical timepoint (n=67, Supplementary Figure 2). We then tested the statistical likelihood of Engel class IA outcome vs. non-Engel class IA outcome in patients with pathogenic Ras/Ras/MAPK variants compared to the rest of the cohort using Fisher's exact test.

Pathway Overrepresentation Analysis

Our initial approach for pathogenic variant discovery relied on variants that were previously reported as pathogenic or likely pathogenic, which yielded an excess of Ras/Raf/MAPK pathway variants. However, to assess the burden of all somatic SNVs with pathogenicity potential—known and unknow—we employed a less biased and comprehensive strategy as follows:

*Creating a High-Quality Call Set*

To minimize the number of false positives and create an accurate set of somatic variants for downstream analyses, we used the paired MuTect2-PON call set, filtered out a sample that had an excessive number of variant calls due to contamination, and applied the following filters to the remaining samples:

1) Variant Allele Frequency (VAF) <0.3
2) Presence in only one individual in the cohort
3) Population allele frequency <0.001 in gnomAD and ExAC
4) Presence on at least 2 alignments with different start positions
5) Supported by minimum 10 ALT reads
6) Minimum 3 ALT reads on forward and reverse orientations
7) Absence from error-prone genomic regions (overlapping RepeatMasker annotations, segmental duplications as annotated in UCSC SegDup, and germline indels)

8) Absence of an adjacent homopolymer repeat with at least 3 units

Filters 4-8 were carried out using our in-house computational pipeline. We independently tested the accuracy of our pipeline using visual inspection of a separate call set using IGV. But to also validate our approach experimentally, we evaluated a random set of variants from the MTLE and control cohorts using amplicon sequencing. All base substitution types were included in this evaluation, but we added more T>A/A>T transversions given an unexpected excess of these variants in the control cohort. For each type of substitution, we drew SNV samples from all the filtered SNVs that are representative of the overall VAF distribution, *i.e.,* since VAF ranges from 0 to 0.3, we broke the range into 6 discrete bins of 0.05 and the number of randomly selected SNVs in each bin was proportional to the number of SNVs in each bin. For the MTLE cohort, 56 out of the 65 tested variants (86.2%) experimentally validated. For the control cohort, 33 out of the 68 variants (48.5%) experimentally validated. However, the excess T>A/A>T transversions in the control cohort, likely introduced during library prep, accounted for 60% of the unvalidated control variants, otherwise the two cohorts had similarly high experimental confirmation rates.

*Pathogenicity Enrichment*

To enrich for somatic SNVs with pathogenicity potential in the high-quality call set, we relied on REVEL, EVE, COSMIC, and SpliceAI annotations. Among the databases, REVEL and SpliceAI provide a pathogenicity score between 0 and 1 for each variant in the database, respectively, and the higher the score, the more likely the variant is to be pathogenic. EVE provides three classifications: “benign”, “pathogenic”, “uncertain”. EVE class80 annotation, which we used, classifies 80% of the variants as pathogenic or benign. All the variants in COSMIC are presumed to be cancer-related and thus pathogenic. Given that the majority (>90%) of the missense variants have REVEL annotations and that only a limited number of variants have EVE and COSMIC annotations, we used the following criteria to enrich for pathogenicity:

1) REVEL score > 0.7 if only annotated by REVEL
2) REVEL score 0.6-0.7 and EVE class80 classification as pathogenic or uncertain, unless present in COSMIC
3) If REVEL score unavailable and EVE class80 classification as pathogenic, unless present in COSMIC
4) SpliceAI score > 0.8

The pathogenicity enriched call sets for the MTLE and control cohorts are shown in Supplementary Table 2.

*Pathway Enrichment Analysis*

To evaluate for the specific biological pathways that may be affected by somatic variants in MTLE, we tested for the overrepresentation of MTLE pathogenicity-enriched somatic SNVs in gene sets curated by the Kyoto Encyclopedia of Genes and Genomes (KEGG). To perform this analysis we used KOBAS[25], an online tool, and selected KEGG pathways as the desired option. Because KOBAS does not take duplicate gene name inputs, all the genes with more than one pathogenicity-enriched somatic variant were counted only once (*e.g.,* PTPN11). We used hypergeometric test/Fisher’s exact test for statistical analysis, and Benjamini and Hochberg (1995) to correct for multiple hypothesis testing in KOBAS.

*Gene Set Overrepresentation Analysis*

To test specifically for over-representation of pathogenicity-enriched somatic variants in specific gets sets, we curated two gene lists: Ras/Raf/MAPK (n=43 genes) and PI3K/Akt/mTOR (n=46 genes) as shown in Table S1. We selected these genes based on our review of other available gene lists (“RAS Pathway v2.0” published on the National Cancer Institute website and “MTOR Signaling Pathway” from the Pathway Interaction Database[26], accessed on April 1, 2022) based on their relevance to neurologic diseases and expression in the brain using the GTEx database. Then we used the hypergeometric test to assess for over-representation of pathogenicity-enriched somatic variants in these gene sets against a background of approximately 19,300 genes targeted by the whole-exome sequencing panel. The pathogenicity-enriched MTLE variants overlapped with 2 Ras/Raf/MAPK genes and 0 PI3K/Akt/mTOR genes out of 13 total genes, whereas in the control call set 0 Ras/Raf/MAPK and PI3K/Akt/mTOR genes were detected out of 34 total genes (n=34). It’s important to note that some of the experimentally confirmed pathogenic Ras/Raf/MAPK variants in the MTLE cohort, were filtered out due to a combination of downsampling and the rigorous filtering that was required for creating a high-quality call set.

Shown below are the specific calculations:

For Ras/Raf/MAPK gene overrepresentation in MTLE:

$$p = 1 - \sum_{i=0}^{1} \frac{\binom{43}{i}\binom{19257}{13-i}}{\binom{19300}{13}} = 4.09 \times 10^{-4}$$

For all other scenarios, the summation term does not exist because there is 0 overlap, so:

$$p = 1$$

We repeated the statistical tests with and without the excess T>A/A>T transversions in the control cohort and confirmed that it did not change the results of this analysis. Since no variants in Ras/Raf/MAPK or PI3K/Akt/mTOR genes were detected in the control cohort, the summation term remained at 0 and p=1.

Experimental Confirmation of Candidate Variants

*Amplicon Validation*

Custom primers were designed for each candidate variant using the default settings in Primer3[1-2] to generate 150-300bp amplicons. The primers were commercially synthesized (IDT) and tested on human genomic DNA (Promega) to confirm generation of only one amplicon product at the expected size. Then 10-50ng of genomic DNA from patients (based on sample availability) were used to create amplicons for sequencing, purified using 2X AMPure XP, and run on a gel for quality control. Amplicons from different samples were pooled together and Illumina sequenced to achieve at least 10,000 reads per each unique amplicon. The raw reads were aligned to the reference genome (hg38) using BWA (version 0.7.17) and the visualized on IGV to confirm the presence of each candidate variant. The variant allele frequencies were calculated based on the total number of REF and ALT alleles. Based on the limitations of PCR amplification and Illumina sequencing, we set the cutoff for confirmation at 0.3% AF.

*ddPCR Validation*

For ddPCR, we ordered commercially available TaqMan primer/probe mixes for BRAF V600E (C_151552107_10, Thermo Fisher) flanking the mutation site. First, we digested 10ng genomic DNA using the HindIII restriction enzyme. Then we used the digested DNA, TaqMan primer/probe mix, and ddPCR Supermix for Probes (Bio-Rad) to perform ddPCR according to the guidelines published in Droplet Digital PCR Applications Guide (Bio-Rad). Finally, we analyzed the ddPCR data using the software QuantSoft (Bio-Rad). The variant allele frequencies were calculated based on the total number of REF and ALT alleles.

Retrospective Review of Somatic Ras/Raf/MAPK and PI3K/Akt/MTOR Variants in Focal Epilepsy

To perform a retrospective review of published lesional focal epilepsy cases with Ras/Raf/MAPK and PI3K/Akt/mTOR somatic variants, we performed a search on pubmed.gov (National Library of Medicine: National Center for Biotechnology Information) using the following criteria:

1) Cases that were published from 2012 through 2022
2) Specific location of the lesion on the brain was identified in the paper
3) Pathogenic somatic variants were identified in Ras/Raf/MAPK and PI3K/Akt/mTOR genes

We used the following search terms in pubmed:

- “FCD” “somatic” “mutation” “mtor”
    - 36 search results
    - 14 papers met the above criteria
- “mutation” “epilepsy” “fcd”
    - 45 search results
    - 3 papers met the above criteria
- “mtor” “focal cortical dysplasia”
    - 139 search results
    - 1 paper met the above criteria
- "ras" "mutation" "ganglioglioma"
    - 6 search results
    - 2 papers met the above criteria

- raf mutation ganglioglioma
    - 90 search results
    - 9 papers met the above criteria
- "braf" "mutation" "dnet"
    - 6 search results
    - 1 paper met the above criteria
- mutation ganglioglioma
    - 12 search results
    - 3 paper met the above criteria
- glioma ras mapk
    - 101 search results
    - 1 paper met the above criteria
- ras mutation focal cortical dysplasia
    - 13 search results
    - 2 papers met the above criteria
- glioma braf mutation
    - 615 search results
    - 1 paper met the above criteria
- focal cortical dysplasia mutation epilepsy
    - 232 search results
    - 1 paper met the above criteria

The required information was extracted from the main body of the paper or supplemental documents and analyzed as follows:

1) All the identified variants and their corresponding lesion locations were curated in two separate tables based on their presence in the Ras/Raf/MAPK or PI3K/Akt/mTOR pathways.
2) Lesions outside the cerebral cortex were excluded from further analysis.
3) Lesion locations were reassigned as frontal, parietal, parieto-occipital, occipital, or temporal based on the available information.

For plotting the cases on the brain in figure 1B:

1) The absolute number of cases for each brain region were normalized based on the total number of Ras/Raf/MAPK or PI3K/Akt/mTOR cases.
2) Then the five brain regions were converted to X,Y coordinates and plotted on the surface of a representative drawing of the brain using ggplot2 in RStudio. The original drawing of the brain was obtained from Biorender.com and modified in Adobe Illustrator.
3) The circle diameters for each point represent the normalized case count for each brain region.

For the bar plot comparing the absolute number of temporal and extra-temporal cases in figure 1B:

1) All the frontal, parietal, parieto-occipital, occipital lesions were regrouped as “extra-temporal”.
2) The binomial test was used for statistical comparison of temporal and extra-temporal cases.

<u>Cell Culture and Plasmid Transfection</u>
KYSE520 (female) cell purchased from Cobioer was cultured in RPMI1640 (Gibco) supplemented with 10% (v/v) FBS (Gibco), 100 units/mL penicillin and 100 mg/mL streptomycin (Gibco). Human HEK293T (female) cell was cultured in DMEM(Gibco) supplemented with 10% (v/v) FBS (Gibco), 100 units/mL penicillin and 100 mg/mL streptomycin (Gibco). All cells were cultured at 37ºC in a humidified atmosphere of 95% air and 5% $CO_2$. Plasmids were transfected into KYSE520 and HEK 293T cells by using jetOPTIMUS (Polyplus, 117-15) according to the manufacturer’s instructions.

<u>Live Cell Imaging</u>
Cells were grown on 24-well glass bottom plate (Cellvis, P24-1.5H-N) and images were obtained with the Leica TCS SP8 confocal microscopy system using a 100x oil objective (NA=1.4) after transfection. Cells were imaged on a heated stage (37℃) by heating chamber and supplemented with warmed (37℃) humidified air. Fluorescent images were processed and assembled into figures using LAS X (Leica) and Fiji.

Fluorescence Recovery After Photobleaching (FRAP)

KYSE520 Cells were grown on 24-well glass bottom plate (Cellvis, P24-1.5H-N) and the FRAP assay was performed using the FRAP module of the Leica TCS SP8 confocal microscopy system. The $Shp2^{mut}$-mEGFP was bleached using a 488-nm laser beam. Bleaching was focused on a circular region of interest (ROI) using 100% laser power and time-lapse images were collected. Fluorescence intensity was measured using Fiji. Background intensity was subtracted, and values are reported relative to pre-bleaching time points. The FRAP results were analyzed by GraphPad Prism6.0.

Protein Expression and Purification

These mutants of Shp2 were generated using standard PCR methods and confirmed by DNA sequencing and were inserted into a pET28a(+) vector with a 6× histidine tag on the N terminus to the constructs. Then, these constructs were transformed into *Escherichia coli* BL21(DE3) cells and grown in LB medium at 37°C to an optical density at OD600 of 0.8. The expression of recombinant proteins was induced by 1mM IPTG at 16℃ for 18h. Cells were harvested by centrifugation at 8000 rpm for 15min at 4℃. After centrifugation, cells were lysed in buffer containing 25mM Tris-HCl pH 8.0, 500mM NaCl and 1mM PMSF, followed by supercentrifugation at 18000 rpm for 30min at 4℃. The supernatant was loaded onto a HisTrap FF chelating column (GE healthcare) in 25 mM Tris-HCl pH 8.0, 500mM NaCl, 1mM DTT and proteins were eluted with the addition of 300mM imidazole. Then, fractions containing Shp2 were concentrated and loaded onto a HiLoad 16/600 Superdex 200 pg column (GE Healthcare) using buffer containing 25mM Tris-HCl pH 8.0, 150mM NaCl, 2mM DTT. Fractions were collected according to the results of SDS-PAGE, and then these proteins were concentrated to 10 mg/mL or more, and stored at -80℃.

*In Vitro* Liquid-Liquid Phase Separation (LLPS) Assay

For the LLPS assay, the purified $Shp2^{wt}$ or $Shp2^{mut}$ protein at 8μM was mixed with a LLPS buffer containing 20mM Tris pH8.0, 10%(w/v) PEG3350 (Sigma) and incubated for 5min at room temperature. Next, 2μL of each sample was pipetted onto a glass dish and imaged using a Leica microscope. Under the same conditions, 30μL of each sample was added into 384-well white polystyrene plate with clear flat bottom, and the value of OD600 was measured by using a Thermo Varioskan™ LUX microplate reader.

Western Blot

Whole cell lysates were prepared in 3 × loading buffer with phosphatase inhibitors (Sigma, P0044 and P5726). Gel electrophoreses of protein lysates were run on 4%–12% NuPAGE Bis-Tris gels (Thermo Fisher, NP0336BOX) at constant voltage of 150V in MES SDS running buffer (Thermo Fisher, NP0002) for 50 min, then transferred to Nitrocellulose membranes using the iBlot gel transfer stack (Thermo Fisher, IB23002), Program 3 for 7 min. Membrane was blocked with 5% (m/v) BSA for 1 hr at room temperature and then incubated with primary antibodies at 4ºC overnight and second antibodies at room temperature for 1hr. The antibodies were used as follow: pERK1/2 (CST#4370), ERK1/2 (CST#9102), SH-PTP2 (Santa Cruz#sc7384), and α/β-tubulin (CST#2148).

Statistical Analysis for Experimental Data

All data are presented as the mean ± standard error of mean (SEM) from independent determinations, and statistical analyses were done using the software Graphpad Prism version 6.0 (GraphPad Software, Inc,; La Jolla, CA, USA). Differences of means were tested for statistical significance with two/one-tailed Student's t-test.

Phosphorylated Erk1/2 Staining of MTLE Tissue

We performed phosphorylated Erk1/2 (pERK1/2) staining on paraffin-embedded archival hippocampal tissue for cases with the following Ras/Raf/MAPK variants: *PTPN11* c.1507G>A (p.G503R), *PTPN11* c.1508G>T (p.G503V), *NF1* c.654+1G>A (altered splicing), *KRAS* c.35G>A (p.G12D). Additionally, we included one case without a known Ras/Raf/MAPK pathogenic variant as control: EP13101. Immunohistochemistry was performed using the pERK1/2 antibody (CST#4370) at 1:500 dilution.

Single cell RNA Sequencing Reanalysis of Human MTLE Hippocampal Tissue

We downloaded the data from a previously published 10X Genomics 3' single cell RNA sequencing (scRNAseq) dataset that used surgical hippocampal resections from MTLE patients[27]. The raw sequencing data for subjects A56 and A57 were downloaded since they best represented the age of subjects in our cohort. Then the expression matrices were generated by CellRanger (10X Genomics, version 5.0.0) using the recommended settings. Low quality cells, as determined by <500 expressed genes and >10% mitochondrial DNA were filtered out. Then we used Seurat V3[28] to perform all the downstream analyses and plotting. Briefly, the expression matrices from the two cases were log-normalized, integrated, dimensionality reduced using the Uniform Manifold Approximation and Projection for Dimension Reduction (UMAP) approach, and clustered. Known gene markers from prior studies[29] were used to identity all the major cell clusters. Finally, the log-normalized RNA expression levels for *PTPN11, KRAS, SOS1, NF1,* and *BRAF* were plotted.

### Supplementary Tables

**Supplementary Table 1. Demographic and Clinical Characteristics of Patients and Neurotypical Controls**

A. Summary of demographic and clinical characteristics of MTLE patients and neurotypical controls.

B. Demographic information, clinical characteristics, tissue availability, recruitment site, 2-year surgical outcome, and sequencing strategy for patients with MTLE.

C. Demographic information, cause of death, tissue availability, source, and sequencing strategy for neurotypical controls.

**Supplementary Table 2. List of Variants Predicted to be Pathogenic**

A. List of all the filtered variants predicted to be pathogenic *in silico* based on REVEL, EVE, and SpliceAI scores in MTLE (A) and control (B) call sets.

**Supplementary Table 3. Curated Gene Sets for Over-representation Analysis**

| **eTable 1. Basic Demographic and Clinical Characteristics** | | |
|---|---|---|
| **Characteristic** | **MTLE Patients (N=105)** | **Neurotypical Controls (N=30)** |
| Age at surgery -- yr | | |
| Mean ± SD | 30.2 ± 13.8 | 30.4 ± 18.0 |
| Median (range) | 32.0 (0.9-66) | 37.0 (7-67) |
| Sex -- no. (%) | | |
| Male | 52 (49.5) | 19 (63.3) |
| Female | 53 (50.5) | 11 (36.7) |
| Histopathologic findings -- no. (%) | | |
| MTS | 91 (86.7) | NA |
| MTS + LEAT | 4 (3.8) | NA |
| MTS + FCD | 2 (1.9) | NA |
| no MTS | 8 (7.6) | NA |
| MRI findings | | |
| MTS | 60 (57.1) | NA |
| no MTS | 15 (14.3) | NA |
| NA | 30 (28.6) | NA |

MTS: Mesial Temporal Sclerosis

no MTS: No evidence of Mesial Temporal Sclerosis

LEAT: Low-Grade Epilepsy-Associated Tumor

FCD: Focal Cortical Dysplasia

NA: Data not available

**Supplementary Table 1A. Summary of demographic and clinical characteristics of MTLE patients and neurotypical controls.**

| Patient ID | Sex | Age (provided as a range, per MerRxiv policy) | Manner of Death | Tissue Sequenced | Recruitment Site | Sequencing Approach |
|---|---|---|---|---|---|---|
| UMB5925 | Female | 6-10 | Natural | Hippocampus<br>Prefrontal neocortex | NIH NeuroBioBank | Exome (IDTERPv1custom) |
| UMB5446 | Female | 16-20 | Natural | Hippocampus<br>Heart | NIH NeuroBioBank | Exome (IDTERPv1custom) |
| UMB5699 | Male | 36-40 | Accidental | Hippocampus<br>Heart | NIH NeuroBioBank | Exome (IDTERPv1custom) |
| UMB5889 | Male | 26-30 | Natural | Hippocampus<br>Muscle | NIH NeuroBioBank | Exome (IDTERPv1custom) |
| UMB5926 | Male | 21-25 | Natural | Hippocampus<br>Muscle | NIH NeuroBioBank | Exome (IDTERPv1custom) |
| UMB5705 | Male | 31-35 | Natural | Hippocampus<br>Heart | NIH NeuroBioBank | Exome (IDTERPv1custom) |
| UMB5387 | Male | 11-15 | Accidental | Hippocampus<br>Heart | NIH NeuroBioBank | Exome (IDTERPv1custom) |
| UMB4782 | Male | 16-20 | Accidental | Hippocampus<br>Heart | NIH NeuroBioBank | Exome (IDTERPv1custom) |
| UMB5958 | Male | 21-25 | Natural | Hippocampus<br>Muscle | NIH NeuroBioBank | Exome (IDTERPv1custom) |
| UMB5630 | Female | 36-40 | Natural | Hippocampus<br>Kidney | NIH NeuroBioBank | Exome (IDTERPv1custom) |
| UMB5629 | Female | 31-35 | Natural | Hippocampus<br>Prefrontal neocortex | NIH NeuroBioBank | Exome (IDTERPv1custom) |
| UMB5566 | Female | 11-15 | Natural | Hippocampus<br>Kidney | NIH NeuroBioBank | Exome (IDTERPv1custom) |
| UMB5818 | Male | 41-45 | Natural | Hippocampus<br>Prefrontal neocortex | NIH NeuroBioBank | Exome (IDTERPv1custom) |
| UMB5609 | Female | 51-55 | Natural | Hippocampus<br>Prefrontal neocortex | NIH NeuroBioBank | Exome (IDTERPv1custom) |
| UMB5919 | Male | 61-65 | Accidental | Hippocampus<br>Prefrontal neocortex | NIH NeuroBioBank | Exome (IDTERPv1custom) |
| UMB5697 | Male | 66-70 | Natural | Hippocampus<br>Heart | NIH NeuroBioBank | Exome (IDTERPv1custom) |
| UMB5540 | Female | 51-55 | Natural | Hippocampus<br>Prefrontal neocortex | NIH NeuroBioBank | Exome (IDTERPv1custom) |
| UMB5895 | Male | 46-60 | Natural | Hippocampus<br>Prefrontal neocortex | NIH NeuroBioBank | Exome (IDTERPv1custom) |
| HCT17-HFK | Female | 41-45 | Natural | Hippocampus<br>Temporal neocortex | NIH NeuroBioBank | Exome (IDTERPv1custom) |
| UMB5875 | Female | 36-40 | Natural | Hippocampus<br>Liver | NIH NeuroBioBank | Exome (IDTERPv1custom) |
| UMB5242 | Male | 11-15 | Natural | Hippocampus<br>Liver | NIH NeuroBioBank | Exome (IDTERPv1custom) |
| UMB5309 | Female | 11-15 | Natural | Hippocampus<br>Liver | NIH NeuroBioBank | Exome (IDTERPv1custom) |
| UMB5643 | Male | 16-20 | Natural | Hippocampus<br>Prefrontal neocortex | NIH NeuroBioBank | Exome (IDTERPv1custom) |
| UMB6055 | Male | 16-20 | Accidental | Hippocampus<br>Liver | NIH NeuroBioBank | Exome (IDTERPv1custom) |
| UMB5551 | Male | 51-55 | Natural | Hippocampus<br>Temporal neocortex | NIH NeuroBioBank | Exome (IDTERPv1custom) |
| CON16001 | Male | 51-55 | Natural | Hippocampus<br>Temporal neocortex | NIH NeuroBioBank | Exome (IDTERPv1custom) |
| UMB5882 | Female | 36-40 | Natural | Hippocampus<br>Prefrontal neocortex | NIH NeuroBioBank | Exome (IDTERPv1custom) |
| UMB5862 | Male | 56-60 | Natural | Hippocampus<br>Prefrontal neocortex | NIH NeuroBioBank | Exome (IDTERPv1custom) |
| UMB13350 | Male | 41-45 | Natural | Hippocampus<br>Temporal neocortex | NIH NeuroBioBank | Exome (IDTERPv1custom) |
| UMB5452 | Male | 66-70 | Natural | Hippocampus<br>Prefrontal neocortex | NIH NeuroBioBank | Exome (IDTERPv1custom) |

**Supplementary Table 1C. Demographic information, cause of death, tissue availability, source, and sequencing strategy for neurotypical controls.**

| eTable 3. Curated Gene Sets for Over-representation Analysis | | | |
|---|---|---|---|
| **Ras/Raf/MAPK Genes** | | **PI3K/Akt/MTOR Genes** | |
| GRB2 | RIN1 | PIK3CA | PLD1 |
| HRAS | RIT1 | AKT3 | PLD2 |
| KRAS | SHOC2 | TSC1 | PDPK1 |
| NRAS | CBL | TSC2 | PRKCA |
| NF1 | DAB2IP | RHEB | PRR5 |
| RRAS | PRKCA | MTOR | PXN |
| RRAS2 | PRKCB | DEPDC5 | RICTOR |
| SOS1 | PRKCE | AKT1 | RPS6KA1 |
| SOS2 | PRKCZ | PTEN | RPS6KB1 |
| PTPN11 | LZTR1 | NPRL2 | RPTOR |
| BRAF | A2ML1 | NPRL3 | SGK1 |
| RAF1 | SPRY2 | AKT1S1 | SREBF1 |
| MAPK1 | SPRY3 | ATG13 | SSPO |
| MAPK3 | RALA | CCNE1 | ULK1 |
| MAP2K1 | NF2 | CDK2 | ULK2 |
| MAP2K2 | CAMK2B | CLIP1 | YWHAB |
| SYNGAP1 | FGFR1 | EIF4A1 | YWHAE |
| RASA1 | FGFR2 | EIF4B | YWHAG |
| RASA2 | FGFR3 | EIF4E | YWHAH |
| RASA3 | | EIF4EBP1 | YWHAQ |
| RASA4 | | DEPTOR | YWHAZ |
| RASAL1 | | DDIT4 | YY1 |
| RASGRF1 | | FBXW11 | |
| RASGRF2 | | MLST8 | |

**Supplementary Table 3. Curated Gene Sets for Over-representation Analysis**

### Supplementary Figures

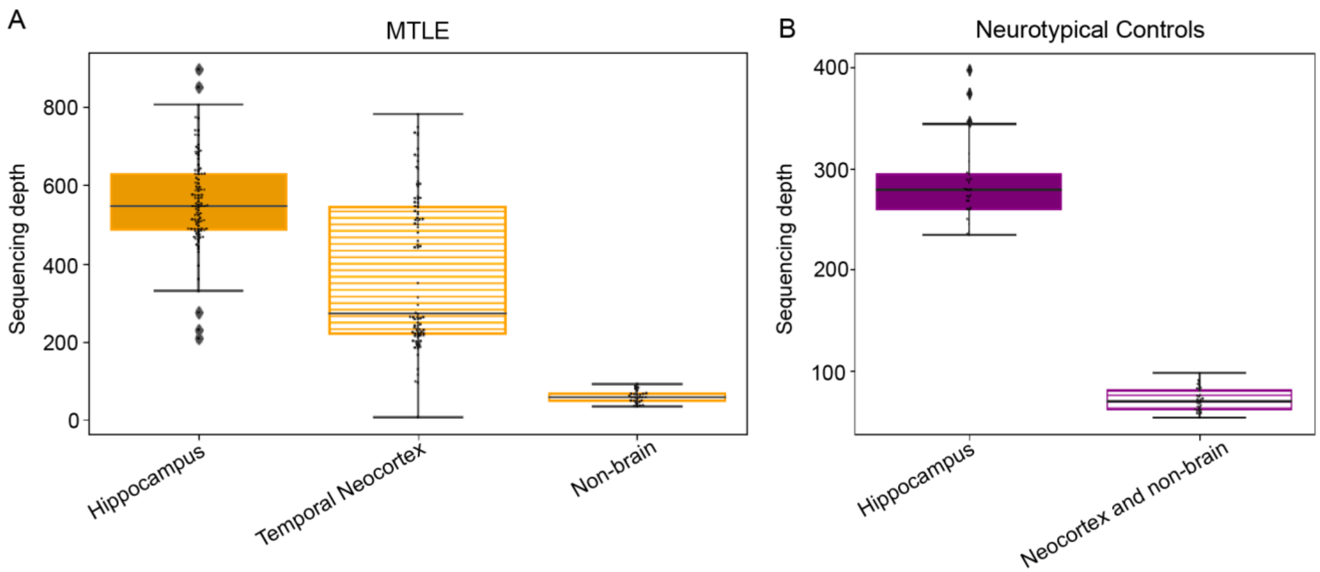

**Supplementary Figure 1. Mean Sequencing Depth for MTLE and Neurotypical Control Samples**
As shown in panel A, >500X mean sequencing depth was achieved for most MTLE hippocampal samples. MTLE temporal neocortical samples were also sequenced deeply, although for paired variant calling, they were downsampled to 50X to be used as germline controls for somatic variant detection, as described in the Supplementary Methods. The sequencing depth for the neurotypical control samples is shown in panel B. For any analyses comparing MTLE and control samples, the MTLE bam files were downsampled to the same average mean depth as the controls, 284X, as described in the Supplementary Methods. Boxes show the lower and upper quartiles and the line inside the box is median. Whiskers indicate lower and upper extremes of the distribution. Each point represents an individual sample.

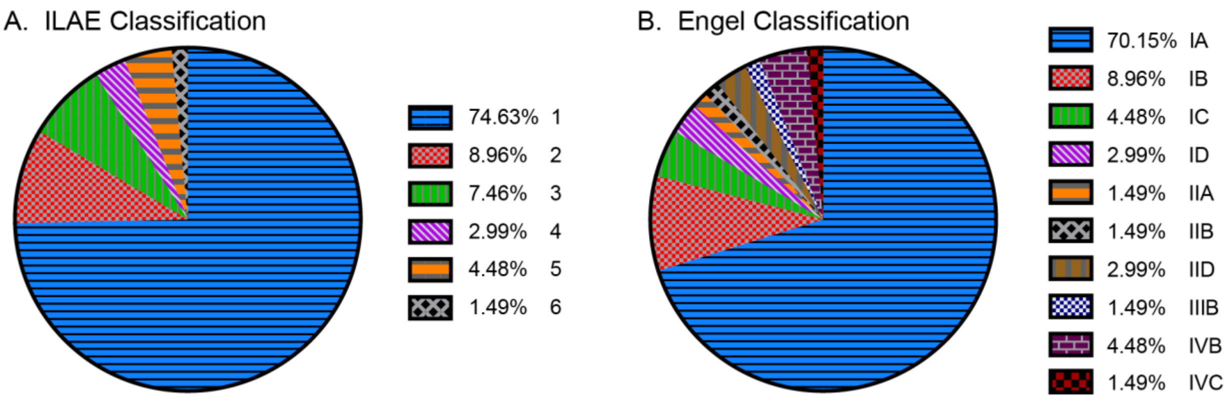

**Supplementary Figure 2. Two-year Epilepsy Surgery Outcome**
The two-year epilepsy surgery outcome for study participants is summarized in pie chart format based on both ILAE and Engel classification systems. Only participants with sufficient clinical follow up data at the 2-year postsurgical timepoint are shown here (n=67).

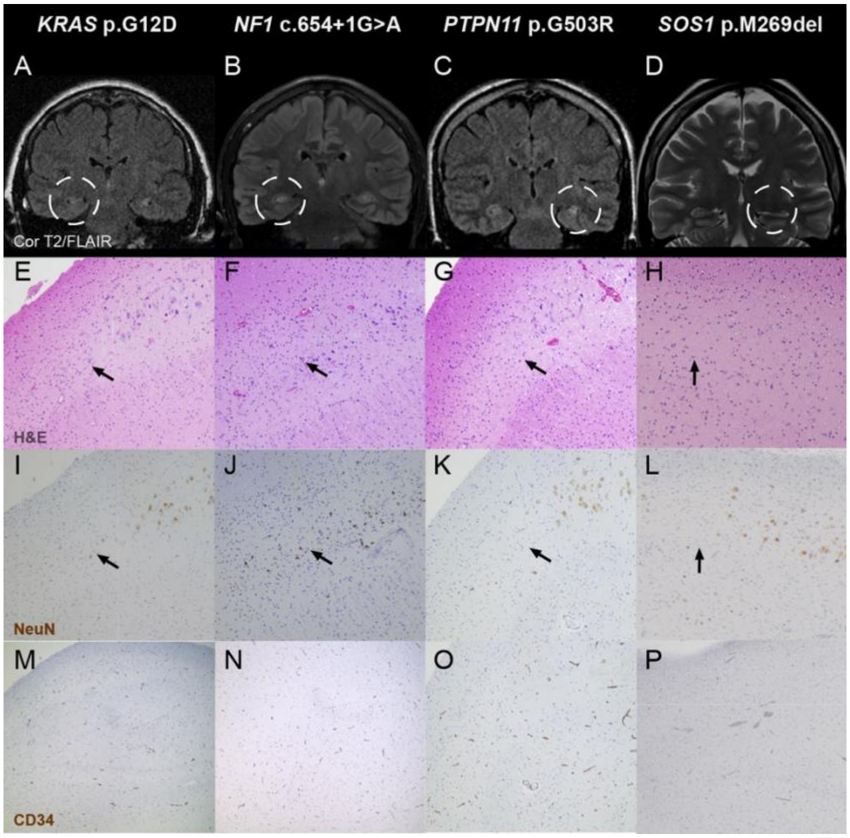

**Supplementary Figure 3. MTLE Cases with Activating Ras/Raf/MAPK Variants and MTS-only Pathology.** Radiographic and histopathologic findings in patients with MTS-only pathology are show in panels A-P. Each column exhibits data for one patient. A-D) Representative T2/FLAIR coronal MRI images show hippocampal atrophy (A-D) and T2/FLAIR signal hyperintensity (B-D), which are consistent with MTS. Expansile or contrast-enhancing lesions are not seen. E-L) Representative histopathological images from the surgically resected hippocampi (demarcated by the dashed white circle in the first row). H&E and NeuN staining show neuronal loss in the Cornu Ammonis (CA) region of the hippocampus (black arrows). M-P) Background-level CD34 staining of blood vessels with no evidence of a glioneuronal tumor. The higher resolution images were obtained at 100-200X and the lower resolution images were obtained at 40X.

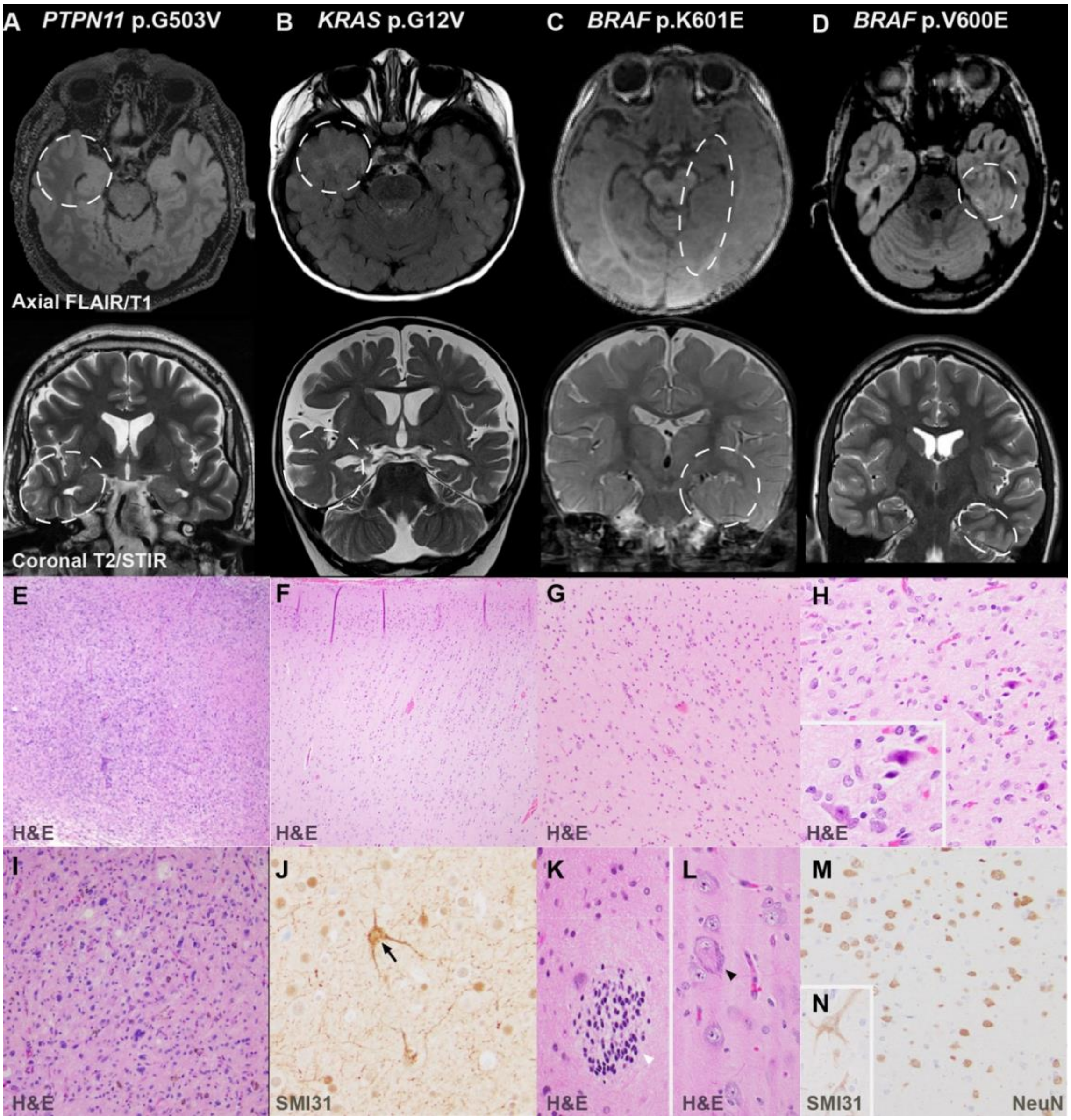

**Supplementary Figure 4. MTLE Cases with Activating Ras/Raf/MAPK Variants and MTS+ Pathology**
Each column in this figure corresponds to one patient. In the first two rows, representative FLAIR/T1 axial and T2/STIR coronal MRI images highlight the main radiographic abnormality in each case (demarcated by the dashed white circle in the first row). In mcdbose315, panel C, the dysplasia starts in the anterior medial temporal region and extends posteriorly to the temporal/occipital junction, indicating an early gestational origin. Rows 3 and 4 show some of the key histopathologic findings in each patient. Areas of hypercellularity shown in panels E and I are consistent with a low-grade tumor. Focal microcolumnar arrangement of cortical neurons in panel F and SMI31+ atypical neurons (black arrow) in panel J are consistent with a diagnosis of FCD type 1A. Panel G shows indistinct cortical layers which in conjunction with hamartias (white arrowhead) in panel K and dysplastic neurons (black arrowhead) in panel L are supportive of a diagnosis of FCD type 2A. Mild glial and neuronal atypia in panel H (highlighted in the inlet) and clusters of neurons with crowding that lack lamination in panel M, are consistent with a diagnosis of a glioneuronal tumor. Panel N highlights rare perikaryal staining of atypical neurons with SMI31. The higher resolution images were obtained at 100-200X and the lower resolution images were obtained at 40X.

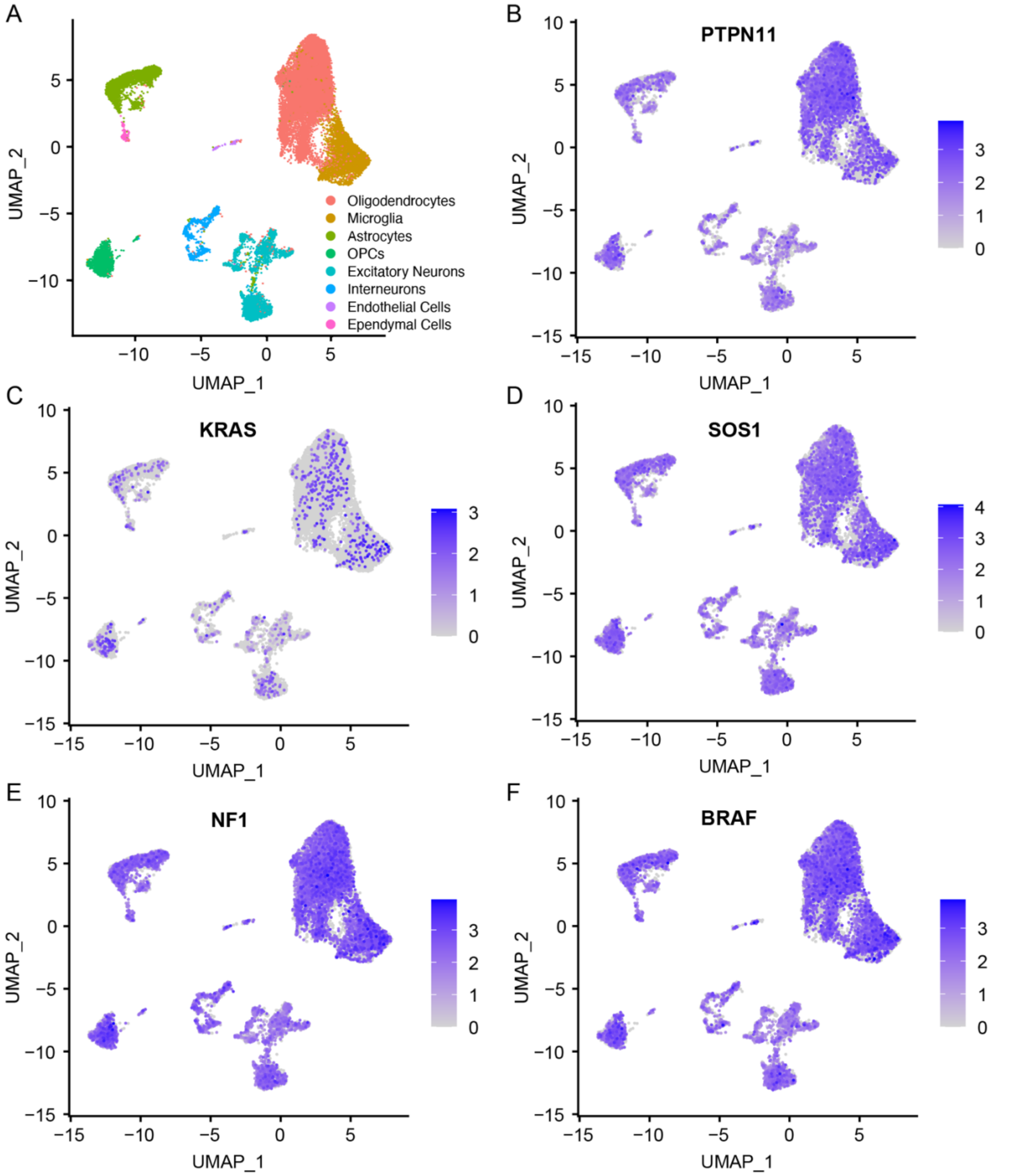

**Supplementary Figure 5. Expression of Ras/Raf/MAPK Genes with MTLE-associated Somatic Variants in the Adult Human Brain**
Published single cell RNA sequencing data from MTLE hippocampal tissue[27] was reanalyzed (Supplementary Figures) to define the major cell clusters (Panel A) and to quantify the RNA expression levels for Ras/Raf/MAPK Genes with MTLE-associate Somatic Variants (Panels B-F). All the Ras/Raf/MAPK genes with pathogenic somatic variants, *PTPN11, KRAS, SOS1, NF1,* and *BRAF,* are expressed in both neurons and glia in the adult human hippocampus from patients with MTLE. The scale bar values represent log-normalized RNA expression levels.

3. Van der Auwera, GA OB. Genomics in the Cloud: Using Docker, GATK, and WDL in Terra. 1st Edition. O'Reilly Media; 2020.

4. Picard Tools - By Broad Institute [Internet]. [cited 2022 May 11]. Available from: http://broadinstitute.github.io/picard/

5. Best practices for data analysis when using UMI adapters to improve variant detection [Internet]. Integrated DNA Technologies. [cited 2022 May 11]. Available from: https://www.idtdna.com/pages/education/videos/detail/best-practices-for-data-analysis-when-using-umi-adapters-to-improve-variant-detection

6. Li H, Handsaker B, Wysoker A, *et al*. The Sequence Alignment/Map format and SAMtools. Bioinformatics. 2009 Aug 15;25(16):2078–2079.

7. Li H. A statistical framework for SNP calling, mutation discovery, association mapping and population genetical parameter estimation from sequencing data. Bioinformatics. 2011 Nov 1;27(21):2987–2993.

8. Smith TS, Heger A, Sudbery I. UMI-tools: Modelling sequencing errors in Unique Molecular Identifiers to improve quantification accuracy. Genome Res. 2017 Jan 18;gr.209601.116.
